# Use of viral load to improve survey estimates of known HIV-positive status and antiretroviral treatment coverage in Kenya

**DOI:** 10.1101/19002592

**Authors:** Peter W. Young, Emily Zielinski-Gutierrez, Joyce Wamicwe, Irene Mukui, Andrea A. Kim, Anthony Waruru, Clement Zeh, Mirjam E. Kretzschmar, Kevin M. De Cock

**Affiliations:** Division of Global HIV & TB, U.S. Centers for Disease Control and Prevention, Nairobi, Kenya; National AIDS & STI Control Programme, Ministry of Health, Kenya; Division of Global HIV & TB, U.S. Centers for Disease Control and Prevention, Atlanta, GA, USA; Julius Center for Health Sciences and Primary Care, University Medical Center Utrecht (UMCU), Utrecht, the Netherlands; Centre for Infectious Disease Control, National Institute of Public Health and the Environment (RIVM), Bilthoven, The Netherlands

**Keywords:** HIV surveillance, antiretroviral treatment, population surveys, biomarkers, Kenya

## Abstract

**Objective:** To compare alternative methods of adjusting self-reported knowledge of HIV-positive status and antiretroviral (ARV) therapy use based on undetectable viral load (UVL) and ARV detection in blood.

**Design:** *Post hoc* analysis of nationally-representative household survey to compare alternative biomarker-based adjustments to population HIV indicators.

**Methods:** We reclassified HIV-positive participants aged 15–64 years in the 2012 Kenya AIDS Indicator Survey (KAIS) that were unaware of their HIV-positive status by self-report as aware and on antiretroviral treatment if either ARVs were detected or viral load was undetectable (<550 copies/mL) on dried blood spots. We compared self-report to adjustments for ARVs measurement, UVL, or both.

**Results:** Treatment coverage among all HIV-positive respondents increased from 31.8% for self-report to 42.5% [95% confidence interval (CI) 37.4–47.8] based on ARV detection alone, to 42.8% (95% CI 37.9–47.8) when ARV-adjusted, 46.2% (95% CI 41.3–51.1) when UVL-adjusted and 48.8% (95% CI 43.9–53.8) when adjusted for either ARV or UVL. Awareness of positive status increased from 46.9% for self-report to 56.2% (95% CI 50.7– 61.6) when ARV-adjusted, 57.5% (95% CI 51.9–63.0) when UVL-adjusted, and 59.8% (95% CI 54.2–65.1) when adjusted for either ARV or UVL.

**Conclusions:** Undetectable viral load, which is routinely measured in surveys, may be a useful adjunct or alternative to ARV detection for adjusting survey estimates of knowledge of HIV status and antiretroviral treatment coverage.

## Introduction

Since the 2007 Kenya AIDS Indicator Survey (KAIS), HIV seroprevalence surveys have often included questions on knowledge of HIV status and antiretroviral (ARV) use among HIV-infected respondents, as well as biomarkers such as viral load (VL) [1–3] and ARV testing. Self-reported knowledge of status and antiretroviral treatment (ART) status can be subject to either positive or negative social desirability bias in some respondents [4] due to the stigma associated with HIV [5,6]. Some respondents may also have inaccurate recall or understanding of detailed questions about their HIV testing and care histories [7].

Antiretroviral testing can be used to adjust self-reported HIV status by reclassifying respondents with ARVs detected in their blood as being previously diagnosed and on ART [8,9]. In the 2012 KAIS 46.9% of HIV-infected respondents self-reported that they were aware of their HIV-positive status, but ARVs were also detected in 21.0% of those not reporting prior HIV diagnosis and 19.3% of those reporting no previous HIV test. However, antiretroviral testing is relatively complex, expensive, and only available within a very limited number of specialized laboratories worldwide, necessitating international shipping, resulting in additional cost, administrative paperwork, and potential for delays.

Unlike ARV testing, which is added exclusively to correct self-reported HIV status and ART use, viral load testing is widely available and routinely included in surveys to estimate population viral suppression (defined as VL < 1000 copies/mL [10]). Undetectable viral load (UVL) is generally indicative of viral suppression due to treatment, hence it could serve as an alternative, indirect marker for treatment. Although the presence of elite controllers (EC) who have UVL in the absence of treatment could confound use of UVL as a proxy for ART use, in US and European cohorts EC are believed to represent only 0.15–1.5% of the HIV-infected population [11], while in East African settings similarly low prevalence of EC has been observed [12,13], limiting the potential impact of this confounding.

Given viral load testing is already conducted routinely in HIV surveys, we examined whether adjusting estimates of knowledge of HIV-positive status and ART coverage using a measure of viral load would achieve similar results to adjustments based on detection of ARVs in a national household survey conducted in Kenya in 2012.

## Methods

The 2012 KAIS included behavioral questions including self-reported HIV and ART status as well as collection of venous blood from which DBS were prepared by field teams and plasma separated and shipped for HIV testing at a national laboratory [2]. After participating in other survey procedures, participants were offered rapid HIV testing by trained HIV counselors in their homes with immediate return of results based on national HIV testing guidelines [14]. Participants testing positive for HIV at the central laboratory were subsequently tested for viral load using the Abbott M2000 platform on DBS subsequently stored at -80°C for future testing. In 2015, DBS were shipped to the University of Cape Town for testing for presence of efavirenz, nevirapine, lopinavir or lamivudine by liquid chromatography tandem mass spectrometry (limit of detection 0.02 µg/mL) [15]. These ARVs were selected to cover first- and second-line regimens in use in Kenya at the time of specimen collection [8,16,17].

We retrospectively re-analyzed survey data to compare self-reported and biomarker-adjusted versions of knowledge of status and ART use among HIV-infected respondents aged 15-64 years.

## Measures

We defined UVL as having a viral load <550 copies/mL on dried blood spots, the limit of detection for the assay used in the study [18]. To calculate UVL-adjusted status, we updated the status for those respondents categorized as ‘unaware’ or ‘aware, not on ART’ with undetectable viral load to ‘aware, on ART’. Similarly, ARV-adjusted status was calculated by updating the status for respondents with ARVs detected in blood to ‘aware, on ART’. For either case, the status for respondents with missing biomarker results was not updated.

We explored differences in self-reported, ARV-adjusted, and UVL-adjusted indicators by age, sex, marital status, educational attainment and mobility. Results were analyzed in R version 3.5.0 [19] using the survey package [20] to adjust and weight results to account for the complex survey design. Wald confidence intervals for survey indicators were calculated on the logarithmic scale and transformed to probability scale using the ‘logit’ method of the svyciprop function in R; confidence intervals previously reported by Kim et al. [21] were calculated on the probability scale.

## Ethical considerations

The 2012 KAIS was approved by the University of California, San Francisco, the U.S. Centers for Disease Control (CDC) in Atlanta, GA, USA and the Kenya Medical Research Institute. Prior to household and individual interviews and blood collection written consent was obtained; in the case of children aged less than 18 years assent was sought in addition to permission from their caregiver or guardian.

## Results

Among 648 HIV-infected respondents, self-reported status was ‘unaware’ among 343 (53.1%), ‘aware, not on ART’ among 100 (15.1%), and ‘aware, on ART’ among 205 (31.8%) (Supplemental Table S1). Of those with UVL and unaware of their HIV-positive status by self-report, 40 also had ARVs detected in blood (Supplemental Table S2). Antiretroviral treatment coverage among all HIV-infected increased from 31.8% (95% CI 27.3–36.6) based on self-report to 42.8% (95% CI 37.9–47.8) when combining self-report and ARV detection, to 46.2% (95% CI 41.3–51.1) when combining self-report and UVL, and finally to 48.8% (95% CI 43.9–53.8) with self-report, UVL or ARVs combined (Table 1). Changes in ART coverage were consistent across demographic characteristics, although the 15–24 year age group saw greater increases when adjusted compared to other age groups (Supplemental Figure S1).

**Table 1.** Self-reported and adjusted estimates of ART coverage among people living with HIV, KAIS 2012

| Characteristic | Self-reported |  |  | ARV only |  |  | Self-report or ARV |  |  | Self-report or UVL (N=648) |  |  | Self-reported, ARV or UVL (N=648) |  |  |
| --- | --- | --- | --- | --- | --- | --- | --- | --- | --- | --- | --- | --- | --- | --- | --- |
|  | (N=648) |  |  | (N=559) |  |  | (N=648) |  |  | UVL (N=648) |  |  | (N=648) |  |  |
|  | n | % | se | n | % | se | n | % | se | n | % | se | n | % | se |
| **Sex** |  |  |  |  |  |  |  |  |  |  |  |  |  |  |  |
| Male | 51 | 27.0 | 3.7 | 64 | 37.8 | 4.7 | 74 | 39.9 | 4.5 | 78 | 41.9 | 4.4 | 84 | 44.3 | 4.5 |
| Female | 154 | 34.7 | 2.5 | 171 | 45.6 | 2.6 | 198 | 44.6 | 2.5 | 217 | 48.8 | 2.5 | 230 | 51.6 | 2.5 |
| **Age group** |  |  |  |  |  |  |  |  |  |  |  |  |  |  |  |
| 15–24 yrs | 6 | 7.1 | 2.7 | 11 | 21.6 | 6.4 | 14 | 20.7 | 5.8 | 19 | 28.8 | 6.2 | 20 | 29.9 | 6.2 |
| 25–34 yrs | 48 | 21.1 | 3.1 | 50 | 25.1 | 3.6 | 65 | 29.3 | 3.5 | 65 | 28.2 | 3.3 | 76 | 33.9 | 3.6 |
| 35–49 yrs | 104 | 41.1 | 3.9 | 121 | 55.0 | 3.8 | 133 | 52.9 | 3.6 | 147 | 58.2 | 3.4 | 152 | 59.5 | 3.3 |
| 50–64 yrs | 47 | 48.0 | 5.4 | 53 | 59.6 | 5.6 | 60 | 60.6 | 5.3 | 64 | 64.0 | 5.0 | 66 | 65.5 | 5.0 |
| **Marital status** |  |  |  |  |  |  |  |  |  |  |  |  |  |  |  |
| Single/never married | 16 | 16.7 | 4.0 | 18 | 24.5 | 5.9 | 22 | 23.8 | 5.1 | 29 | 32.5 | 5.6 | 31 | 33.9 | 5.7 |
| Married/cohabitating | 118 | 31.4 | 3.2 | 141 | 42.0 | 3.4 | 163 | 43.3 | 3.2 | 175 | 46.1 | 3.1 | 186 | 48.6 | 3.2 |
| Divorced /sep /<br>widowed | 71 | 41.5 | 4.0 | 76 | 53.1 | 4.3 | 87 | 52.6 | 4.0 | 91 | 54.2 | 3.9 | 97 | 58.1 | 4.0 |
| **Highest educational attainment** |  |  |  |  |  |  |  |  |  |  |  |  |  |  |  |
| None | 28 | 33.2 | 5.8 | 31 | 35.5 | 5.9 | 39 | 44.6 | 6.0 | 39 | 45.2 | 6.0 | 41 | 46.5 | 6.0 |
| Primary | 94 | 26.7 | 2.8 | 111 | 39.5 | 3.6 | 126 | 38.4 | 3.3 | 137 | 40.8 | 3.3 | 148 | 44.3 | 3.5 |
| Secondary | 10 | 31.4 | 10.2 | 14 | 45.7 | 10.1 | 15 | 43.7 | 9.8 | 16 | 50.7 | 9.3 | 18 | 54.9 | 9.6 |
| Higher | 73 | 40.8 | 4.3 | 78 | 50.6 | 5.0 | 91 | 50.2 | 4.4 | 102 | 55.8 | 4.2 | 106 | 57.2 | 4.1 |
| **Employment** |  |  |  |  |  |  |  |  |  |  |  |  |  |  |  |
| unemployed | 82 | 36.2 | 4.3 | 91 | 50.0 | 4.6 | 106 | 47.6 | 4.3 | 114 | 50.6 | 4.3 | 118 | 52.2 | 4.2 |
| employed | 123 | 29.6 | 2.6 | 144 | 39.0 | 2.8 | 166 | 40.5 | 2.8 | 181 | 44.0 | 2.7 | 196 | 47.2 | 2.8 |
| **Mobility** |  |  |  |  |  |  |  |  |  |  |  |  |  |  |  |
| Not away for >1<br>month in last year | 112 | 31.2 | 3.0 | 126 | 42.0 | 3.5 | 146 | 42.2 | 3.2 | 156 | 44.5 | 3.3 | 166 | 46.8 | 3.2 |
| Away >1 month in last<br>year | 87 | 32.1 | 3.4 | 101 | 43.7 | 4.0 | 117 | 43.7 | 3.6 | 129 | 48.0 | 3.7 | 137 | 51.4 | 3.9 |
| **Total** | 205 | 31.8 | 2.4 | 235 | 42.5 | 2.6 | 272 | 42.8 | 2.5 | 295 | 46.2 | 2.5 | 314 | 48.8 | 2.5 |

Note: self-reported: self-report only, ARV-only: based on presence/absence of ARVs only, self-report or ARV: either self-reported known- positive/on ART or ARVs detected, self-report or UVL: either self-reported known-positive/on ART or viral load was undetectable, self- report, ARV or UVL: either self-reported known-positive/on ART, ARVs detected, or UVL, N: unweighted denominator, n: unweighted numerator, se: standard error. Missing biomarker results were treated as biomarker not present. Percentages and standard errors are weighted and adjusted to account for the survey design.

Knowledge of status increased from 46.9% (95% CI 41.4–52.4) based on self-report to 56.2% (95% CI 50.7–61.6) when adjusting with ARVs, to 57.5% (95% CI 51.9–63.0) when adjusting for UVL, and to 59.8% (95% CI 54.2–65.1) when adjusting for either ARV or UVL (Table 2). Similar to population ART coverage, ART among those with known HIV-positive status also increased from self-report to adjustment, with similar increases between adjustment methods. The youngest age group also saw the biggest impact of adjustment versus self-report for these indicators in both relative and absolute terms.

**Table 2.**
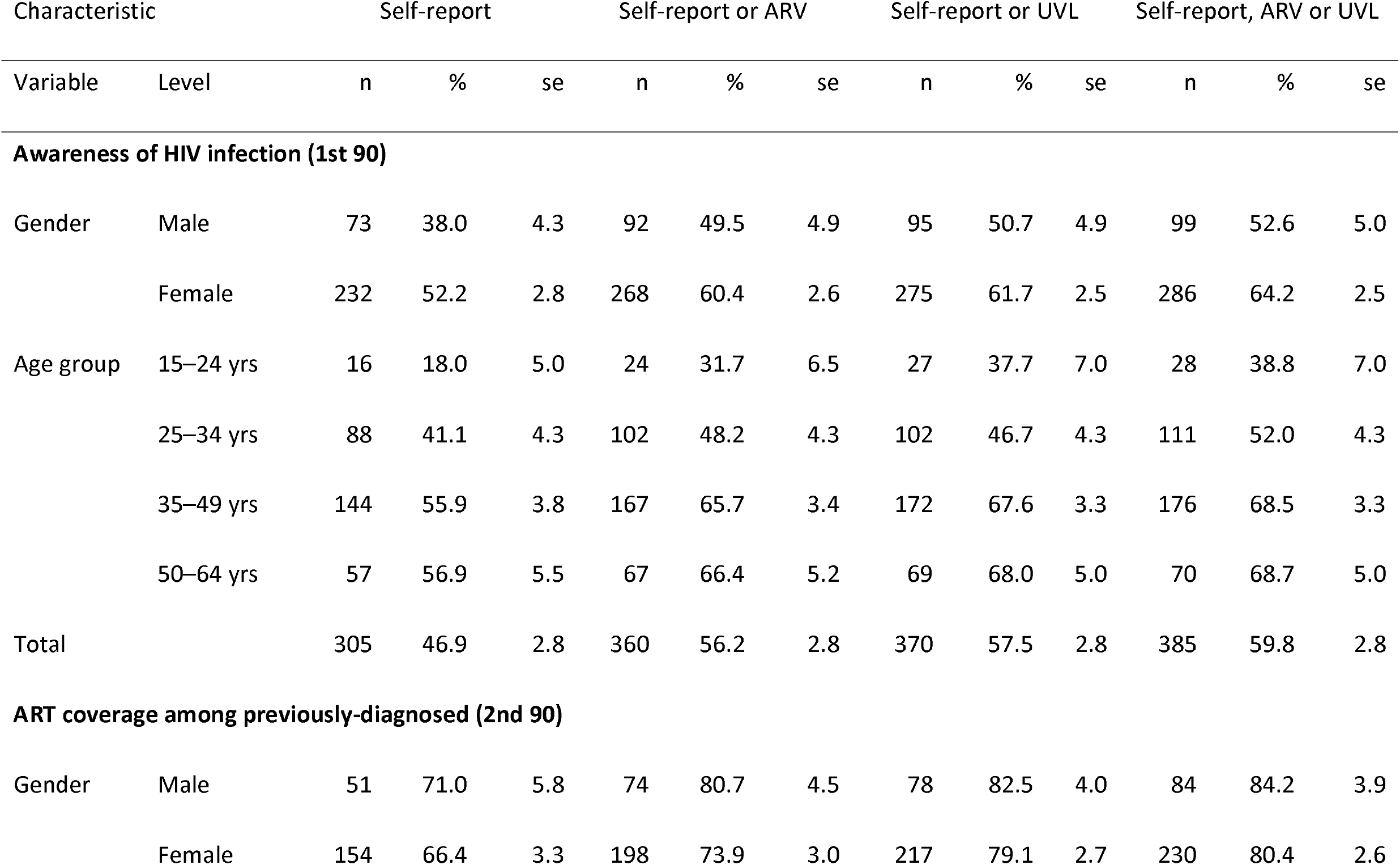
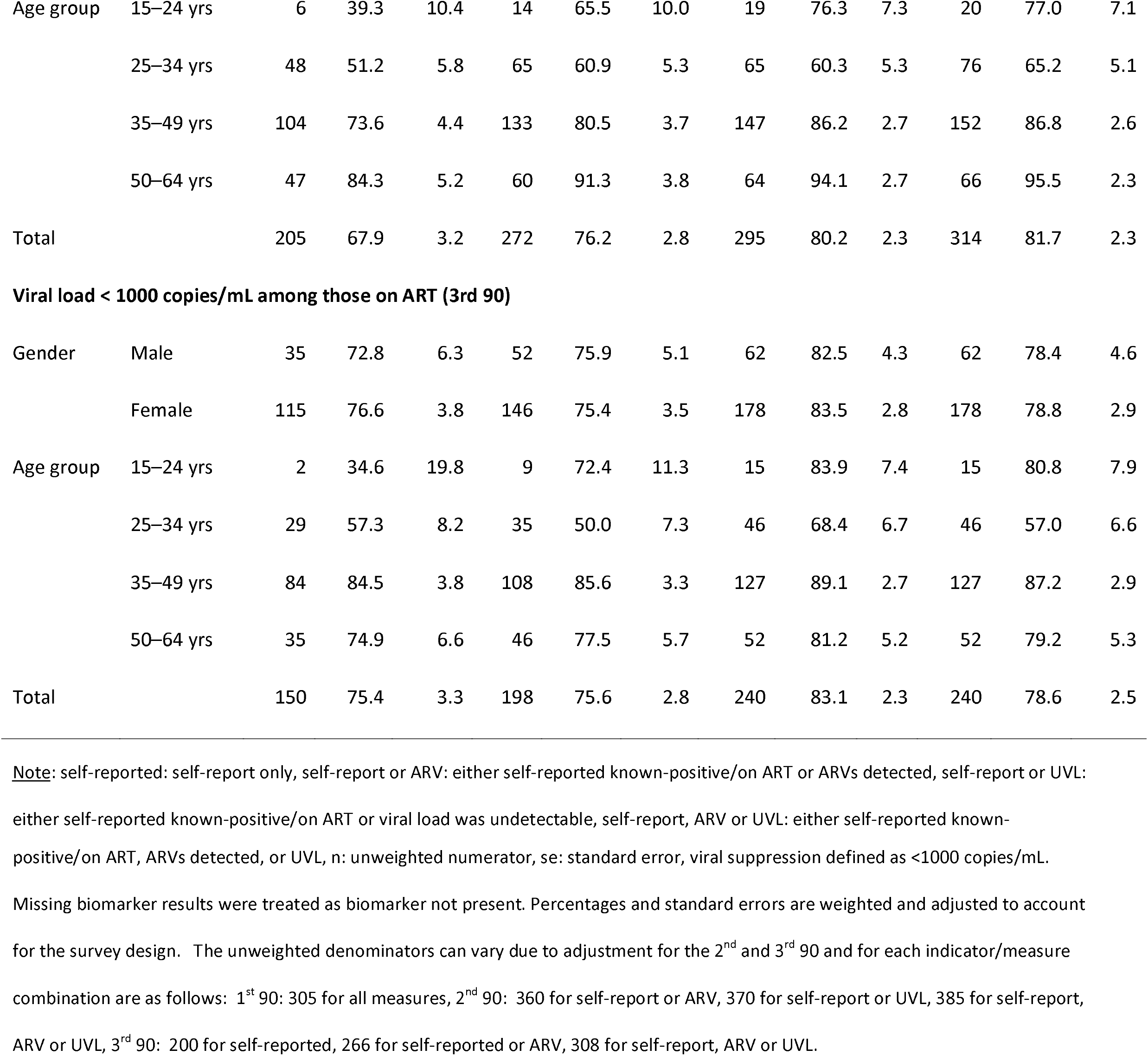
Self-reported and adjusted estimates of HIV care cascade, KAIS 2012

| Characteristic |  | Self-report |  |  | Self-report or ARV |  |  | Self-report or UVL |  |  | Self-report, ARV or UVL |  |  |
| --- | --- | --- | --- | --- | --- | --- | --- | --- | --- | --- | --- | --- | --- |
| Variable | Level | n | % | se | n | % | se | n | % | se | n | % | se |
| **Awareness of HIV infection (1st 90)** |  |  |  |  |  |  |  |  |  |  |  |  |  |
| Gender | Male | 73 | 38.0 | 4.3 | 92 | 49.5 | 4.9 | 95 | 50.7 | 4.9 | 99 | 52.6 | 5.0 |
|  | Female | 232 | 52.2 | 2.8 | 268 | 60.4 | 2.6 | 275 | 61.7 | 2.5 | 286 | 64.2 | 2.5 |
| Age group | 15–24 yrs | 16 | 18.0 | 5.0 | 24 | 31.7 | 6.5 | 27 | 37.7 | 7.0 | 28 | 38.8 | 7.0 |
|  | 25–34 yrs | 88 | 41.1 | 4.3 | 102 | 48.2 | 4.3 | 102 | 46.7 | 4.3 | 111 | 52.0 | 4.3 |
|  | 35–49 yrs | 144 | 55.9 | 3.8 | 167 | 65.7 | 3.4 | 172 | 67.6 | 3.3 | 176 | 68.5 | 3.3 |
|  | 50–64 yrs | 57 | 56.9 | 5.5 | 67 | 66.4 | 5.2 | 69 | 68.0 | 5.0 | 70 | 68.7 | 5.0 |
| Total |  | 305 | 46.9 | 2.8 | 360 | 56.2 | 2.8 | 370 | 57.5 | 2.8 | 385 | 59.8 | 2.8 |
| **ART coverage among previously-diagnosed (2nd 90)** |  |  |  |  |  |  |  |  |  |  |  |  |  |
| Gender | Male | 51 | 71.0 | 5.8 | 74 | 80.7 | 4.5 | 78 | 82.5 | 4.0 | 84 | 84.2 | 3.9 |
|  | Female | 154 | 66.4 | 3.3 | 198 | 73.9 | 3.0 | 217 | 79.1 | 2.7 | 230 | 80.4 | 2.6 |
| Age group | 15–24 yrs | 6 | 39.3 | 10.4 | 14 | 65.5 | 10.0 | 19 | 76.3 | 7.3 | 20 | 77.0 | 7.1 |
|  | 25–34 yrs | 48 | 51.2 | 5.8 | 65 | 60.9 | 5.3 | 65 | 60.3 | 5.3 | 76 | 65.2 | 5.1 |
|  | 35–49 yrs | 104 | 73.6 | 4.4 | 133 | 80.5 | 3.7 | 147 | 86.2 | 2.7 | 152 | 86.8 | 2.6 |
|  | 50–64 yrs | 47 | 84.3 | 5.2 | 60 | 91.3 | 3.8 | 64 | 94.1 | 2.7 | 66 | 95.5 | 2.3 |
| Total |  | 205 | 67.9 | 3.2 | 272 | 76.2 | 2.8 | 295 | 80.2 | 2.3 | 314 | 81.7 | 2.3 |
| **Viral load < 1000 copies/mL among those on ART (3rd 90)** |  |  |  |  |  |  |  |  |  |  |  |  |  |
| Gender | Male | 35 | 72.8 | 6.3 | 52 | 75.9 | 5.1 | 62 | 82.5 | 4.3 | 62 | 78.4 | 4.6 |
|  | Female | 115 | 76.6 | 3.8 | 146 | 75.4 | 3.5 | 178 | 83.5 | 2.8 | 178 | 78.8 | 2.9 |
| Age group | 15–24 yrs | 2 | 34.6 | 19.8 | 9 | 72.4 | 11.3 | 15 | 83.9 | 7.4 | 15 | 80.8 | 7.9 |
|  | 25–34 yrs | 29 | 57.3 | 8.2 | 35 | 50.0 | 7.3 | 46 | 68.4 | 6.7 | 46 | 57.0 | 6.6 |
|  | 35–49 yrs | 84 | 84.5 | 3.8 | 108 | 85.6 | 3.3 | 127 | 89.1 | 2.7 | 127 | 87.2 | 2.9 |
|  | 50–64 yrs | 35 | 74.9 | 6.6 | 46 | 77.5 | 5.7 | 52 | 81.2 | 5.2 | 52 | 79.2 | 5.3 |
| Total |  | 150 | 75.4 | 3.3 | 198 | 75.6 | 2.8 | 240 | 83.1 | 2.3 | 240 | 78.6 | 2.5 |

Note: self-reported: self-report only, self-report or ARV: either self-reported known-positive/on ART or ARVs detected, self-report or UVL:
either self-reported known-positive/on ART or viral load was undetectable, self-report, ARV or UVL: either self-reported known-
positive/on ART, ARVs detected, or UVL, n: unweighted numerator, se: standard error, viral suppression defined as <1000 copies/mL. Missing biomarker results were treated as biomarker not present. Percentages and standard errors are weighted and adjusted to account for the survey design. The unweighted denominators can vary due to adjustment for the 2<sup>nd</sup> and 3<sup>rd</sup> 90 and for each indicator/measure combination are as follows: 1<sup>st</sup> 90: 305 for all measures, 2<sup>nd</sup> 90: 360 for self-report or ARV, 370 for self-report or UVL, 385 for self-report, ARV or UVL, 3<sup>rd</sup> 90: 200 for self-reported, 266 for self-reported or ARV, 308 for self-report, ARV or UVL.

We repeated the analysis excluding the respondents for whom either the ARV or UVL biomarkers were not available; findings were similar (Supplemental Table S3).

## Discussion

In order to balance resources between finding undiagnosed HIV infections, linking patients to HIV treatment, and ensuring retention and adherence to care it is necessary to obtain the best possible estimates of knowledge of HIV-positive status and ART use. We set out to establish whether viral load, a routinely-available marker in HIV surveys, can be used to adjust self-reported estimates of knowledge of HIV-positive status and ART use. In KAIS 2012, UVL-adjusted point estimates were similar to, but slightly greater than ARV-adjusted estimates of knowledge of status and ART coverage, suggesting adjustment with UVL might have been sufficient. When measuring ART coverage, all of the adjusted estimates (ARV only, UVL only, and either ARV or UVL) had overlapping confidence intervals, but are notably higher than estimates based on self-report alone.

The change in estimates when adjusting by ARVs and UVL were similar across demographic groups, but 15–24 year olds did see a larger additional increase when adjusting by UVL. This may indicate poor recent adherence in this group leading to non-detection of the ARV biomarker but undetectable viral load (<550 copies/mL in this study). Li et al found that 37% of patients still had a viral load <200 copies/mL four weeks after interrupting ART [22]. Many ARVs reach undetectable levels in blood within several days of treatment interruption [8,15,23], thus in populations with poor adherence or high rates of treatment interruption, adjusting based on UVL may result in higher estimated ART coverage than measures incorporating ARV detection.

The performance of UVL for adjusting ART use will depend on the prevalence of UVL in the population on HIV treatment. In populations with effective ART programs with high rates of viral suppression in the treated population, it may be a relatively sensitive marker for ART use; however, in populations with poor treatment outcomes a larger proportion of patients on treatment would not have UVL.

The prevalence of elite controllers has not been established in Kenya, hence it is not possible to quantify their influence on the UVL-adjusted estimates, but given the similarity between UVL-adjusted and ARV-adjusted estimates, their potential impact was limited. Simultaneously adjusting for either UVL or presence of ARVs may in fact be closest to true population prevalence of the indicators of interest. Without better data on prevalence of elite controllers in this population it is more conservative to use one or the other marker rather than both combined. In settings with ample evidence of low prevalence of elite control, or where population high ART coverage and immediate treatment initiation means even elite controllers are likely to be on treatment, using the combined indicator would likely represent the most sensitive approach to estimating population-based knowledge of status and ART coverage.

This analysis was subject to several limitations. While adjusting for biomarkers associated with ARV exposure from a single time-point can account for misreporting of status among those on ART, it cannot account for those who misreport their knowledge of HIV-positive status but are not currently on treatment, or those who may be on treatment but transiently non-adherent to medications. Other established methods for reducing bias in self-reported estimates, such as computer-assisted self-interview methods, may also be helpful [24]. This analysis was based on data from a single country with low ART coverage (43.5%) and viral suppression among those on treatment (73.9%) at the time of the survey compared with current program coverage; the UVL adjustment may perform differently in other populations. Simulation or replication of this analysis in a diverse set of populations, including the more recent population-based HIV impact assessments conducted in many countries, could help elucidate the performance of UVL adjustment in different settings. Finally, poor specimen quality could result in false-negative results for both biomarkers. In spite of these limitations, this analysis does strongly suggest that use of UVL to adjust self-reported HIV status and ART use should be considered, especially in surveys where the inclusion of the ARV biomarker may be cost-prohibitive or subject to delays.

## Conclusion

Streamlining the estimation of key HIV program indicators should allow governments, donors and other stakeholders to assess program performance more quickly and affordably. Viral load, which is routinely measured in HIV surveys, may be a useful biomarker for adjusting self-reported indicators of HIV diagnosis and treatment in cross-sectional surveys in absence of, or in addition to, adjustment based on detected ARVs in blood.

## Data Availability

Data for KAIS 2012 are available upon request from NASCOP by emailing, or from the Kenya National Bureau of Statistics by requesting at http://statistics.knbs.or.ke/nada/index.php

## Acknowledgements

We would like to thank the University of Cape Town Department of Clinical Pharmacology for conducting the ART biomarker testing, the National HIV Reference Laboratory for conducting the KAIS 2012 viral load testing, the study teams that collected data in the field, and finally the survey participants.

## Authors’ contributions

PWY, EZG and KDC conceived the study. PWY conducted the analyses. All authors contributed to drafting and critical review of the manuscript.

## Additional files

Additional file 1. Supplementary tables and figures

Word document containing additional supplementary analyses referenced in text.

